# Mechanical thrombectomy vs intravenous rt-PA in medium vessel occlusions: real-world data from the Italian SITS registry

**DOI:** 10.1101/2025.09.11.25335613

**Authors:** M Farè, E Bianchi, G Benina, G Lucchi, M Mercenari, F Innocenti, S Spinelli, G Isgrò, R Orofino, M Galimberti, E Pedranzini, G Orsani, A Giglio, F Pedrazzini, F Pasini, D Montisano, F Santangelo, N Rifino, M Viganò, C Balducci, E Agostoni, R Altavilla, S Amarù, B Ardito, A Baldi, L Bartolomei, M Bellucci, G Bigliardi, G Boero, R Bombardi, I Bosone, G Calabrese, L Caputi, P Cardinali, A Cascio Rizzo, A Cavallini, R Cavallo, P Cerrato, A Chiti, E Coco, L Concari, P Del Dotto, M Del Sette, M Dell’Acqua, D Ferrandi, C Ferrarese, M Filippi, S Forlivesi, M Galletti, A Gasparro, M Gentile, L Godi, M Guidotti, P Invernizzi, N Iorio, P Julita, M Longoni, V Lucivero, M Magoni, F Marchioretto, A Martignoni, G Martusciello, E Medici, P Meineri, V Melas, S Meletti, F Melis, M Melis, G Monteforte, C Mundi, R Napoletano, S Novello, C Paci, C Palmieri, M Paolucci, A Pasquinucci, M Pennisi, S Piffer, V Pinto, M Plocco, A Plutino, M Rasura, A Reia, G Rinaldi, P Rizzo, M Rossi, L Roveri, F Sallustio, A Salmaggi, A Sanna, M Savarese, G Scaglione, M Sessa, L Sicurella, E Spina, S Strumia, R Tarletti, R Tassi, T Tassinari, G Torgano, A Trebbastoni, L Vandelli, M Vista, A Zini, D Toni, S Beretta

**Author notes:** **Corresponding author:** Matteo Farè, Department of Neurology, Fondazione IRCCS San Gerardo dei Tintori, Via G. B. Pergolesi 33, 20900 Monza, Italy.

## Abstract

**Background:** Recent randomized trials have questioned the benefit of endovascular therapy (EVT) for medium vessel occlusion (MeVO) stroke, but data from clinical practice are limited. This study aimed to assess the effectiveness and safety of EVT, with or without intravenous thrombolysis (IVT), versus IVT alone in MeVO stroke using registry-based real-world data.

**Methods:** This retrospective multicenter study included patients from 82 Italian centers in the SITS registry (January 2020–December 2023). Adults with acute ischemic stroke due to MeVO (ACA A1/A2, MCA M2/M3 or more distal, or PCA P1/P2), treated with IVT or EVT±IVT, and with available 90-day modified Rankin Scale (mRS) scores were included. Patients with tandem occlusions were excluded. Propensity score matching (1:1) was used to balance baseline variables. Primary outcome was functional independence (mRS 0–2) at 90 days. Secondary outcomes included in-hospital mortality, intracranial hemorrhage incidence, and recanalization status.

**Results:** Among 1375 total patients, 780 were included and matched (390 per group) by propensity score. Baseline characteristics were balanced. Functional independence at 90 days was achieved in 57.7% of EVT±IVT patients versus 59.2% in the IVT-only group (OR 0.939, 95% CI 0.706–1.248, p=0.663). In-hospital mortality was non significantly lower in the EVT±IVT group (5.4% vs 8.7%, p=0.069). Symptomatic intracranial hemorrhage rates were comparable between groups, although overall hemorrhagic complications were higher with EVT (18.4% vs 11.2%, p<0.0001). Successful recanalization occurred in 81.0% of EVT cases. Stratified analyses by stroke severity and treatment timing showed consistent lack of benefit across all subgroups (all interaction p-values >0.05).

**Conclusions:** EVT did not improve long-term functional outcomes compared to IVT alone in MeVO stroke but was associated with higher hemorrhagic risk. These findings support a cautious approach to EVT in this setting, in line with recent trial evidence.

## Introduction

Acute ischemic stroke is the main neurological disease needing emergency care, especially since revascularization therapies were introduced.^1^ The highest mortality and disabling outcome of ischemic stroke is associated with large-vessel occlusions (LVO), such as intracranial internal carotid artery and M1 segment of middle cerebral artery occlusions.^2^ On the other hand, medium-vessel occlusions (MeVO) usually bear an intrinsic better outcome compared to LVO, since the area of cerebral tissue affected by the hypoperfusion is smaller and can benefit from a larger collateral network.^3^ However, despite the intrinsic better scenario, most patients with ischemic stroke attributed to MeVO do not reach a satisfiable outcome, with over 30% of them not regaining functional independence at 3 months after the event.^3^

Intravenous thrombolysis (IVT) shows clearer efficacy in MeVO than LVO, given the smaller size of the thrombus, despite an early recanalization rate not exceeding 50%.^4^ Endovascular mechanical thrombectomy (EVT) provides evidence-based benefit in LVO ischemic strokes.^5^ However, the utility of EVT in MeVO has been derived from trials which focused on the overall benefit of the procedure, often not taking vessel size into account, and other observational studies did not show significant effectiveness compared to IVT.^6–8^ Two large randomized controlled trials, ESCAPE-MeVO and DISTAL, were published in 2025 and brought new evidence on this topic, both demonstrating no reduction in disability and mortality with EVT compared with best medical treatment, which included IVT if appropriate.^9,10^

This study, SITS-MeVO, aims to assess the benefit of performing thrombectomy, in addition to standard medical therapy, compared to IVT alone, on MeVO stroke outcomes in a routine care setting, using real-world data from the Italian centers participating in the Safe Implementation of Treatments in Stroke (SITS) registry.

## Materials and methods

The study was designed as a retrospective, multicenter, registry-based study, in which we included consecutive patients enrolled between January 1st, 2020, and December 31st, 2023, who met the following criteria: (1) age ≥ 18 years; (2) acute ischemic stroke due to MeVO, defined as per registry options as occlusion in anterior cerebral artery (ACA) A1/A2, middle cerebral artery (MCA) M2/M3 or more distal, or posterior cerebral artery (PCA) P1/P2 segments on CT angiography; (3) treatment with IVT or EVT±IVT; (4) absence of tandem occlusions; (5) availability of 90-day modified Rankin Scale (mRS) score. As the SITS registry does not differentiate between dominant and non-dominant M2 branches, all M2 occlusions were included regardless of dominance pattern; also, the registry categorizes M3 and M4 occlusions together under “M3 or more distal”.

The primary outcome was functional independence at 90 days, defined as mRS score 0–2 in the two treatment groups. Secondary outcomes included in-hospital mortality in the two treatment groups and safety endpoints which included in-hospital and overall mortality, occurrence of symptomatic intracranial hemorrhage according to SITS Monitoring Study (SITS-MOST) criteria^11^, and recanalization status when available.

From the initial cohort of 1375 patients meeting inclusion criteria, 444 (32.3%) had undergone EVT. Among these, 54 patients were excluded due to missing data on primary outcome or key matching variables, resulting in 390 EVT patients. To minimize confounding and selection bias, treatment groups were compared using a matched cohort design: a propensity score was calculated using a logistic regression model, with treatment (EVT±IVT vs. IVT only) as dependent variable and baseline demographic and clinical variables which were considered as possible confounders as independent variables. Possible confounders were the following: age, baseline NIH Stroke Scale (NIHSS) score, vascular risk factors, onset-to-treatment times, imaging features, such as dense artery sign, site and side of the occlusion. Patients were matched 1:1 by both site of occlusion and propensity score using nearest-neighbor matching without replacement.

The final analyses were conducted on the matched population only. Descriptive statistics on the main demographic and clinical variables were performed in the entire sample and comparing the two treatment groups. Categorical variables were reported as counts and percentages, continuous variables as medians with interquartile ranges. Group comparisons were performed using chi-square or Fisher’s exact tests for categorical variables and Wilcoxon-Mann-Whitney test for continuous variables. The odds ratio (OR) for good clinical outcome was estimated using logistic regression models with treatment group as the independent variable and treatment group as dependent variable. A 90-days survival analysis comparing the two treatment groups was also performed with Kaplan-Meier survival curves and Log-rank test. Stratified analyses of the primary outcome by NIHSS categories (<5, 5-14, >14) and onset-to-treatment time (≤180 minutes, >180 minutes) were performed, testing first treatment-by-covariate interactions to assess the presence of significant differences in the effect of treatment between strata.

All tests were two-tailed with a 5% level of significance and analyses were carried out with the SAS software (version 9.4, SAS Institute, Cary, NC, USA).

## Results

A total of 1375 patients with MeVO stroke met the inclusion criteria and were retrieved from the SITS database. Of these, 444 (32.3%) underwent EVT, while 931 (67.7%) received IVT alone. After excluding patients with missing data on primary outcome or matching variables (Figure 1), 780 patients were included in the final matched analysis, with 390 patients in each treatment group (EVT±IVT versus IVT alone). In the EVT±IVT group 150 patients received both treatments, while 240 received only mechanical thrombectomy. The patients excluded from the matching process (n = 595, 43.3%) differed significantly from the matched cohort (Table S1): they were slightly older (median age 79 vs 76 years, p<0.0001), had lower baseline NIHSS scores (median 7 vs 10, p<0.0001) and higher baseline disability (mRS 3-5, 9.75% vs 4.74%, p=0.0005), and were predominantly treated with IVT alone (90.9% vs 205 50.0%).

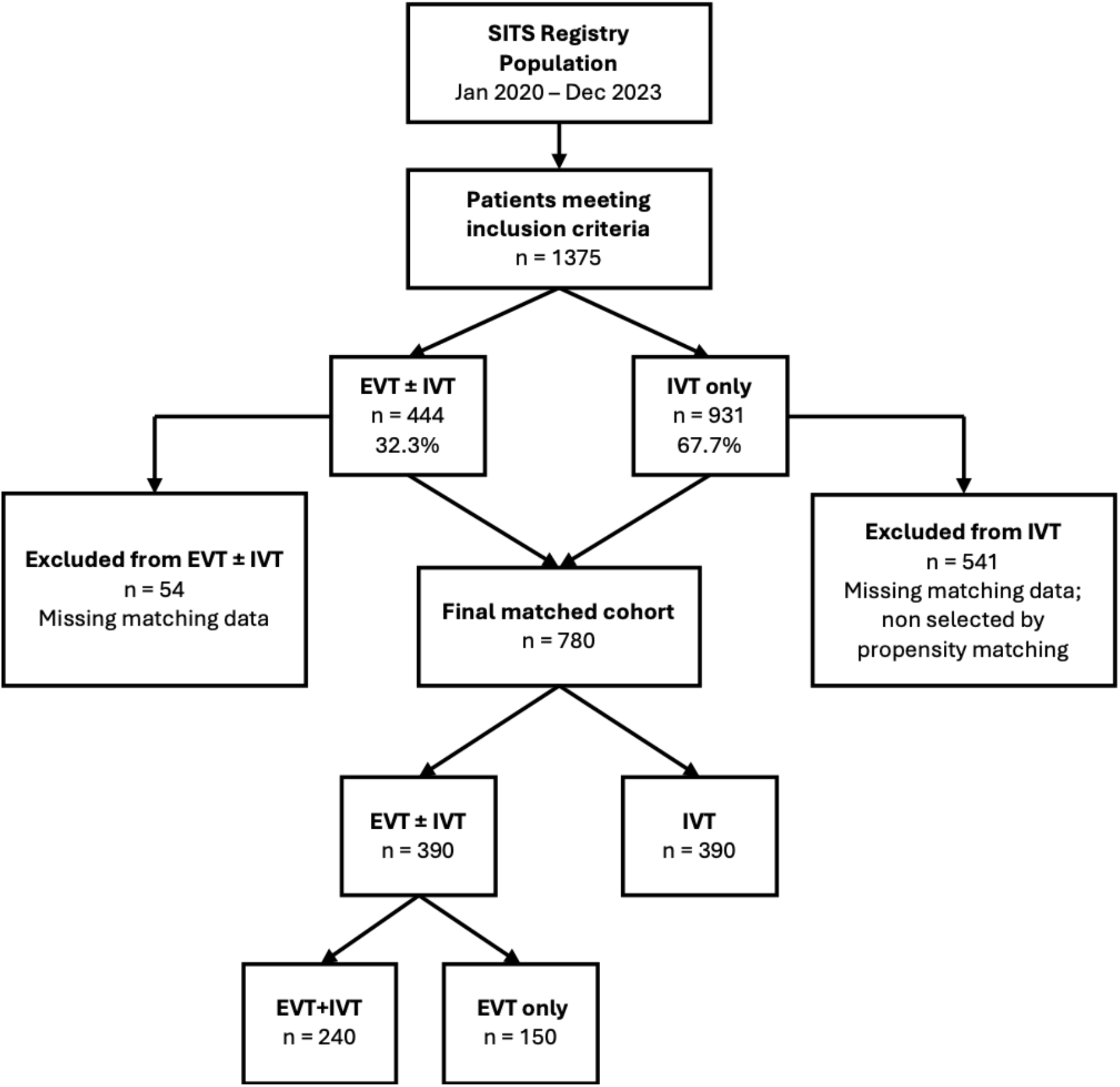

### Baseline Characteristics

The baseline characteristics were well-balanced between groups following propensity score matching (Table 1). The median age was 76 years (interquartile range 67-83), with no significant difference between the EVT±IVT group (78 years, IQR 68-83) and the IVT-only group (75 years, IQR 67-83; p=0.277). Male patients comprised approximately half of each group (49.7% vs 50.0%, p=0.943). Most patients had favorable baseline functional status, with 95.3% having a pre-stroke mRS score of 0-2 213 (p=0.613).

**Table 1.** Population baseline characteristics. **Abbreviations:** EVT, endovascular therapy; IVT, intravenous thrombolysis; mRS, modified Rankin Scale; NIHSS, National Institutes of Health Stroke Scale. ^a^Data only applicable for patients who underwent EVT+IVT

| Characteristic | Total (n = 780) | EVT ± IVT (n = 390) | IVT (n = 390) |  |
| --- | --- | --- | --- | --- |
|  | n (%) or median (Q1-Q3) | n (%) or median (Q1-Q3) | n (%) or median (Q1-Q3) | p-value |
| **Demographics** |  |  |  |  |
| Age, years | 76 (67-83) | 78 (68-83) | 75 (67-83) | 0.277 |
| Sex, male | 389 (49.87) | 194 (49.74) | 195 (50.00) | 0.943 |
| Baseline mRS |  |  |  | 0.613 |
| 0 - 2 | 743 (95.26) | 370 (94.87) | 373 (95.64) |  |
| 3 - 5 | 37 (4.74) | 20 (5.13) | 17 (4.36) |  |
| **Risk Factors** |  |  |  |  |
| Hypertension | 560 (71.79) | 286 (73.33) | 274 (70.26) | 0.340 |
| Diabetes Mellitus | 131 (16.79) | 68 (17.44) | 63 (16.15) | 0.632 |
| Hyperlipidemia | 296 (37.95) | 151 (38.72) | 145 (37.18) | 0.658 |
| Smoking, currently | 90 (12.33) | 45 (12.26) | 45 (12.40) | 0.954 |
| Atrial Fibrillation | 204 (26.15) | 114 (29.23) | 90 (23.08) | 0.051 |
| Previous Stroke | 71 (9.10) | 37 (9.49) | 34 (8.72) | 0.709 |
| **Clinical Presentation** |  |  |  |  |
| NIHSS Score on admission | 10 (6-15) | 10 (6-15) | 9 (5-15) | 0.271 |
| Median onset to needle time, minutes | 155 (118-215) | 155 (118-210)<sup>a</sup> | 155 (119-215) | 0.368 |
| Median onset to groin time, minutes | / | 265 (200-385) | / | / |
| **Imaging** |  |  |  |  |
| Dense artery sign | 384 (49.23) | 192 (49.23) | 192 (49.23) | 0.966 |
| Occlusion Site |  |  |  | 1.000 |
| M2 | 690 (88.46) | 345 (88.46) | 345 (88.46) |  |
| M3 or more distal | 18 (2.31) | 9 (2.31) | 9 (2.31) |  |
| A1 or A2 | 12 (1.54) | 6 (1.54) | 6 (1.54) |  |
| P1 or P2 | 60 (7.69) | 30 (7.69) | 30 (7.69) |  |
| Occlusion Side |  |  |  | 0.560 |
| Right | 320 (41.03) | 164 (42.05) | 156 (40.00) |  |
| Left | 460 (58.97) | 226 (57.95) | 234 (60) |  |

Vascular risk factors were similarly distributed between groups, including hypertension (73.3% vs 70.3%, p=0.340), diabetes mellitus (17.4% vs 16.2%, p=0.632), current smoking habit (12.3% vs 12.4%, p=0.954), and hyperlipidemia (38.7% vs 37.2%, p=0.658). Atrial fibrillation showed a higher prevalence in the EVT±IVT group compared to the IVT-only group (29.2% vs 23.1%, p=0.051).

Clinical presentation was comparable between groups, with median NIHSS scores of 10 (IQR 6-15) in the EVT±IVT group and 9 (IQR 5-15) in the IVT-only group (p=0.271). Median onset-to-needle time was similar between groups at 155 minutes (IQR 118-210 vs 119-215, p=0.368). For patients receiving endovascular therapy, the median onset-to-groin time was 265 minutes (IQR 200-385).

The distribution of occlusion sites was identical between groups due to matching criteria, with M2 segment occlusions representing the vast majority of cases (88.5%), followed by posterior circulation occlusions in P1 or P2 segments (7.7%). Left-sided occlusions were more frequent than right-sided 225 occlusions (59.0% vs 41.0%, p=0.560).

### Primary outcome

Functional independence at 90 days (mRS 0-2) was achieved in 57.7% of patients in the EVT±IVT group compared to 59.2% in the IVT-only group (Table 2). This difference was not statistically significant (odds ratio 0.939, 95% confidence interval 0.706-1.248, p=0.663), indicating no superiority of EVT over IVT alone for MeVO stroke.

**Table 2.** modified Rankin Scale 0-2 at 90 days. **Abbreviations:** EVT, endovascular therapy; IVT, intravenous thrombolysis; mRS, modified Rankin Scale; OR, odds ratio; CI, confidence interval.

|  | Total (n = 780) | EVT ± IVT (n = 390) | IVT (n = 390) |  |  |
| --- | --- | --- | --- | --- | --- |
|  | n (%) | n (%) | n (%) | OR (95% CI) | p-value |
| **mRS 0-2** | 456 (58.46) | 225 (57.69) | 231 (59.23) | 0.939 (0.706 – 1.248) | 0.663 |

### Secondary outcomes

Secondary outcome analysis results are listed in Table 3. In-hospital mortality occurred in 5.4% of patients in the EVT±IVT group compared to 8.7% in the IVT-only group, showing a trend towards lower mortality with EVT, which approached statistical significance (p=0.069).

**Table 3.** Secondary outcomes. **Abbreviations:** EVT, endovascular therapy; IVT, intravenous thrombolysis; TICI, thrombolysis in cerebral infarction; sICH, spontaneous intracranial hemorrhage; HI1, Hemorrhagic Infarction type 1; HI2, Hemorrhagic Infarction type 2; PH1, Parenchymal Hemorrhage type 1; PH2, Parenchymal Hemorrhage type 2. ^a^defined as mRS 6 at discharge; ^b^defined as TICI 2b or 3; ^c^missing data from 192 subjects; ^d^missing data from 221 subjects; ^e^according to SITS-MOST definition.

|  | Total (n = 780) | EVT ± IVT (n = 390) | IVT (n = 390) |  |
| --- | --- | --- | --- | --- |
|  | n (%) | n (%) | n (%) | p-value |
| ***In-hospital mortality<sup>a</sup>*** | 55 (7.07) | 21 (5.40) | 34 (8.74) | 0.069 |
| ***Successful Recanalization<sup>b</sup>*** |  | 179 (81.00) |  |  |
| ***TICI score<sup>c</sup>*** |  |  |  |  |
| 0 |  | 21 (9.50) |  |  |
| 1 |  | 3 (1.36) |  |  |
| 2a |  | 18 (6.34) |  |  |
| 2b |  | 61 (25.00) |  |  |
| 3 |  | 118 (53.39) |  |  |
| ***Revascularization attempts<sup>d</sup>*** |  |  |  |  |
| 1 |  | 117 (59.39) |  |  |
| 2 |  | 44 (22.34) |  |  |
| > 3 |  | 36 (18.29) |  |  |
| ***Ischemic lesion size at 24h CT*** |  |  |  | 0.415 |
| Total | 9 (3.50) | 4 (2.99) | 5 (4.07) |  |
| 2/3 of the vascular territory | 68 (26.46) | 40 (29.85) | 28 (22.76) |  |
| 1/3 of the vascular territory | 180 (70.04) | 90 (67.16) | 90 (73.17) |  |
| ***Hemorrhage at 24 h CT<sup>e</sup>*** |  |  |  | < 0.0001 |
| None | 636 (85.14) | 310 (81.58) | 326 (88.83) |  |
| HI1 | 43 (5.76) | 31 (8.16) | 12 (3.27) |  |
| HI2 | 27 (3.61) | 14 (3.68) | 13 (3.54) |  |
| PH1 | 27 (3.61) | 15 (3.95) | 12 (3.27) |  |
| PH2 | 14 (1.87) | 10 (2.63) | 4 (1.09) |  |

The safety profile differed significantly between treatment groups, particularly regarding intracranial hemorrhage rates. While 11.2% of patients in the IVT-only group experienced an intracranial hemorrhage according to SITS-MOST classification, this proportion was higher in the EVT±IVT group at 18.4% (p<0.0001). As expected, contrast extravasation occurred exclusively in the EVT±IVT group (5.3% vs 0%). However, the rates of more severe hemorrhagic complications, such as parenchymal hemorrhage types 1 and 2 (PH1, PH2), were similar between the two groups, respectively 6.6% in EVT±IVT and 4.4% in IVT (p = 0.20; OR 1.54, 95%CI 0.81-2.94), indicating that the excess hemorrhagic risk was primarily limited to less severe forms.

Ischemic lesion size on 24-hour CT imaging, defined as proportion of total vascular territory, showed no significant difference between treatment groups (p=0.415), with most patients in both groups demonstrating infarcts involving one-third or less of the vascular territory (67.2% in EVT±IVT vs 73.2% in IVT-only).

Among patients who underwent endovascular procedures, data on TICI score and the number of revascularization attempts were available in only 198/390 (50.8%) and 169/390 (43.3%) of cases, respectively. In the subset with available data, successful recanalization (TICI 2b–3) was achieved in 81.0% of patients.

Kaplan-Meier survival analysis (Figure 2) demonstrated a trend toward improved cumulative survival rates with EVT±IVT compared to IVT alone over the 3-month follow-up period (p=0.0675), consistent with the lower in-hospital mortality observed in the EVT group (5.4% vs 8.7%, p=0.069). However, this survival benefit did not reach statistical significance and was not evident when analyzing the EVT-only (p=0.2928) or EVT+IVT (p=0.1083) subgroups separately, likely due to reduced statistical power in these subgroups.

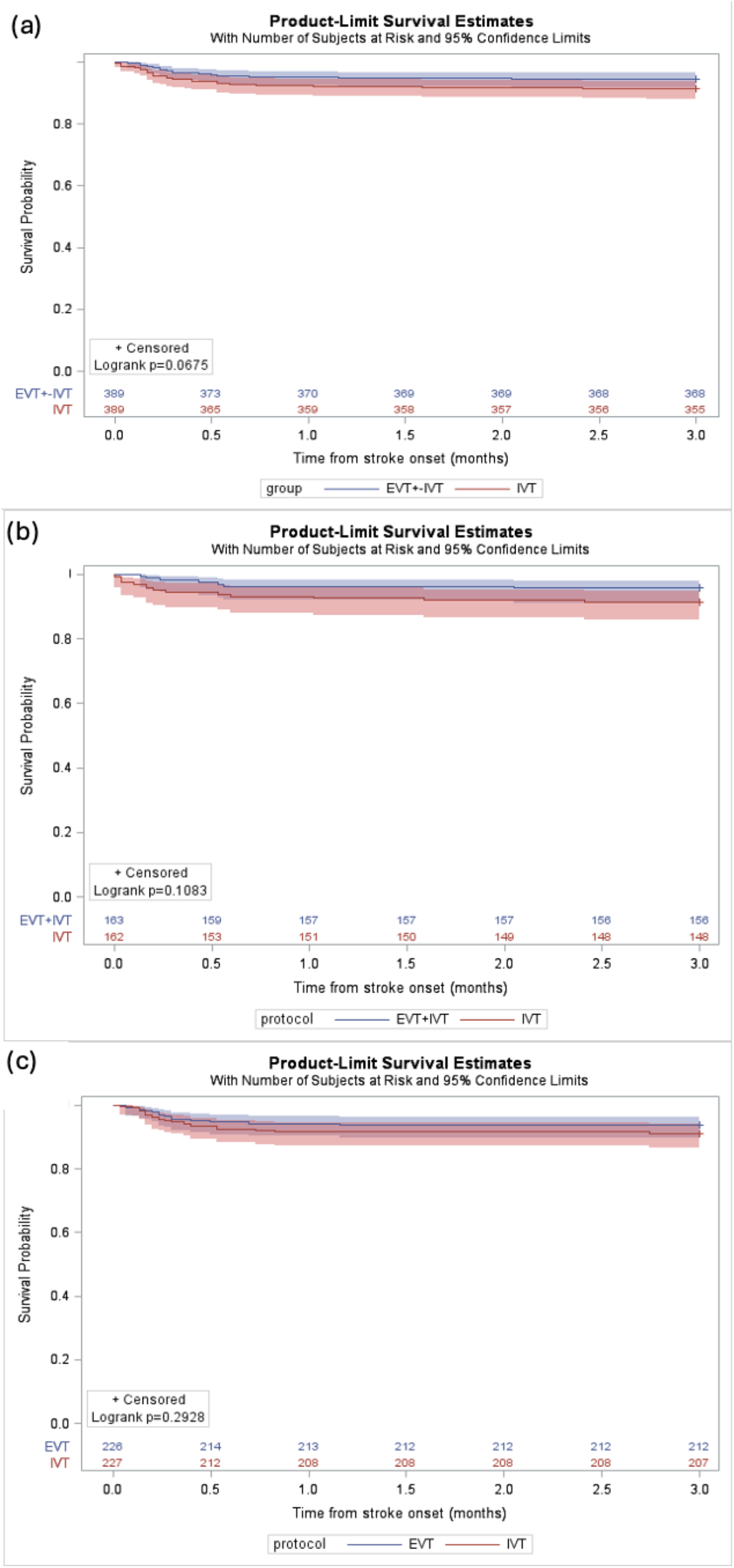

### Stratified Analysis

Interaction testing revealed no significant treatment-by-NIHSS interaction (p=0.218) or treatment-by-onset-to-treatment-time interaction (p=0.148), indicating that the treatment effect did not vary significantly across severity or timing strata. In the descriptive stratified analysis by NIHSS categories (Table S2), functional independence rates in the EVT±IVT versus IVT-only groups were: 79.1% versus 83.3% for NIHSS <5 (OR 0.79, 95% CI 0.30-2.06, p=0.628); 65.1% versus 62.2% for NIHSS 5-14 (OR 1.14, 95% CI 0.77-1.68, p=0.506); and 34.9% versus 36.3% for NIHSS >14 (OR 0.94, 95% CI 0.54-1.64, p=0.832). When stratified by onset-to-treatment time (Table S2), functional independence rates were: 66.1% versus 58.3% for ≤180 minutes (OR 1.39, 95% CI 0.75-2.57, p=0.290); and 56.5% versus 61.8% for >180 minutes (OR 0.80, 95% CI 0.52-1.23, p=0.311). None of these stratum-specific comparisons reached statistical significance, consistent with the overall findings of no treatment benefit.

## Discussion

This multicenter, real-world study based on the SITS registry from Italian centers, indicates that EVT, with or without IVT, does not provide superior functional outcomes compared to IVT alone in patients with MeVO stroke. These findings align with the recently published results from the ESCAPE-MeVO and DISTAL randomized controlled trials, which similarly demonstrated no reduction in disability and mortality with thrombectomy compared to best medical treatment in this patient population.^9,10^ While our study employed a different methodological approach as a registry-based observational retrospective study with propensity score matching rather than randomized assignment, the convergence of results across different study designs strengthens the evidence base regarding the limited benefit of routine endovascular therapy for MeVO. The consistency of findings across randomized trials and registry data suggests that the absence of benefit is robust across different patient populations and clinical settings.^12–14^
Several methodological strengths enhance the reliability of these findings. The multicenter design across 82 Italian SITS centers provide substantial external validity and generalizability to routine clinical practice. The use of propensity score matching with careful attention to occlusion site matching helped minimize selection bias inherent in observational studies, creating comparable treatment groups despite the non-randomized design. The SITS registry infrastructure ensures standardized data collection protocols across participating centers, maintaining consistency in outcome definitions and follow-up procedures that strengthen the validity of cross-center comparisons.

A particularly important finding of this study is the slightly higher rate of intracranial hemorrhage in the EVT group, which was primarily due to an increase in minor events. More importantly, rates of more severe hemorrhages (PH1, PH2) were comparable between groups. This suggests that while EVT carries a higher hemorrhagic burden, the excess risk is mostly limited to less severe forms, which nonetheless warrant careful consideration, given the absence of functional benefit.

Conversely, we observed a non-significant trend toward higher in-hospital mortality in the IVT-only group (8.7% vs 5.4%, p = 0.069): although this may be influenced by residual confounding, it raises the possibility that some patients not offered EVT might have experienced fatal complications due to persistent occlusion. This underscores the need to identify clinical or imaging profiles that could help select high-risk patients who might still benefit from thrombectomy on a case-by-case basis.

The absence of significant treatment-by-covariate interactions confirms that the lack of EVT benefit is consistent across different stroke severities and treatment timing windows. This homogeneity strengthens our primary findings and indicates that the absence of functional benefit applies broadly across MeVO patients typically considered for endovascular therapy. Even in patients with severe strokes or across different treatment timings, no EVT advantage was observed, supporting the clinical interpretation that routine EVT does not provide superior outcomes compared to IVT alone in MeVO stroke. Importantly, this stratified analysis represents a novel contribution that extends beyond the findings of recent randomized trials, ESCAPE-MeVO and DISTAL, providing additional granular evidence that the lack of EVT benefit is consistent across clinically relevant patient subgroups, thereby enhancing the external validity and clinical applicability of the evidence base for MeVO management.

Taken together, these findings support a cautious approach to routine EVT in MeVO. While current evidence does not justify its widespread use, clinical judgment should be prioritized, and thrombectomy may still be considered in specific scenarios, such as outside the thrombolysis time window if the imaging profile (eg. high penumbra/core ratio) is favorable. As EVT device technology continues to evolve rapidly, the risk-benefit profile may change, warranting ongoing surveillance and re-evaluation of outcomes in future clinical studies. The composition of our cohort provides important insights into routine care MeVO management patterns. Only 31.5% of eligible patients underwent EVT, with the remainder receiving IVT alone. This distribution likely reflects the gradual adoption of endovascular therapy for medium vessel occlusions during our study period (2020-2023), as well as the more conservative patient selection criteria historically applied to MeVO compared to LVO. The significant differences between matched and non-matched patients, particularly the age older than > 80 years, higher Rankin scale at baseline, and milder stroke severity in the excluded group, suggest that our results primarily apply to patients younger than 80 years old with moderate to severe MeVO strokes and better baseline functional status, who have traditionally been considered better candidates for EVT.

Our study has limitations to be considered. First, its retrospective nature introduces potential for unmeasured confounding, despite the use of propensity score matching. Second, the sample size, though comparable to other RCTs, was constrained by incomplete data in the original pre-matching cohort of 1375 patients.

Third, the matching process excluded 595 patients (43.3%) from the analysis. Only 444 (32.3%) had undergone EVT, reflecting the selective and evolving use of EVT for MeVO during the study period. Excluded patients were older, had milder strokes (lower NIHSS), and higher baseline disability, and were predominantly treated with IVT alone. This suggests a selection bias toward younger and/or more severely affected patients, limiting generalizability to elderly or mildly affected populations who are now increasingly considered for EVT.

Fourth, the classification of MeVO was limited by the anatomical definitions available in the SITS registry. Inclusion of A1 and P1 segments, sometimes categorized as large vessels^15^, and lack of differentiation between proximal and distal M2 occlusions may have led to partial misclassification. However, comparable NIHSS scores (median 10 vs 9) suggest overall balance in stroke severity. Moreover, the registry does not distinguish between dominant and non-dominant M2 branches, where dominant M2 occlusions are often not considered MeVO in many trials and observational studies. Given that 82% of our cohort underwent propensity score matching, this lack of anatomical specificity regarding M2 dominance patterns represents a source of potential misclassification that could affect the interpretation of our results.

Fifth, missing data affected the assessment of procedural outcomes: TICI scores and revascularization attempts were available in only ∼50% and 43% of EVT cases, respectively. Moreover, unlike recent RCTs, our dataset included only revascularized patients, lacking a best medical treatment control group. Lastly, infarct size thresholds were applied uniformly across vascular territories, although validation differs by territory^16^, possibly introducing minor classification bias.

The predominance of M2 occlusions in our cohort strengthens the validity of our findings for this subgroup, but limits generalizability to less represented territories, such as A1/A2, P1/P2, and M3. These sites may differ in anatomy and collateral patterns, potentially influencing EVT response. Additionally, our study spanned a period of evolving endovascular techniques and included the early COVID-19 pandemic, which likely impacted workflows and treatment decisions. These temporal and technical factors should be considered when interpreting outcome variability and the gradual adoption of EVT for MeVO.

Building on our findings and the alignment with recent RCTs, several clinical and research considerations emerge. The absence of functional benefit from EVT, combined with increased hemorrhagic risk, supports a conservative stance in future guideline updates. EVT should not be routinely recommended for MeVO stroke but considered selectively, especially when IVT is contraindicated or imaging suggests salvageable tissue.

Even among patients with moderate severity and good baseline status, who were more likely to receive EVT, no benefit emerged. This reinforces the need for refined selection algorithms integrating vessel anatomy, perfusion imaging, and individualized risk–benefit assessment. Given the resource intensity and lack of outcome improvement, EVT for MeVO may not be cost-effective at a population level. Targeted use in subgroups with clear potential for benefit is essential to justify procedural costs and avoid overtreatment.

The findings of this study suggest important opportunities for expanded investigation across the broader international SITS network: world-wide analysis could address several critical questions that remain unanswered, including the impact of healthcare system variations on treatment selection and outcomes, geographic differences in clinical practice patterns, and the identification of specific patient subgroups who might derive benefit from endovascular therapy. Such an expanded dataset would provide substantially greater statistical power to detect clinically meaningful differences in rare but important outcomes and could enable more sophisticated analyses of treatment effect heterogeneity across different patient characteristics and clinical presentations.

## Conclusion

This multicenter, retrospective, real-world study did not show a functional benefit of EVT over IVT alone in MeVO stroke. Our results align with recent randomized trials, reinforcing the need for a cautious, individualized approach. Future research should focus on identifying specific clinical and imaging characteristics that might predict treatment response in specific subgroups, while continued technological advancement may eventually alter the therapeutic landscape for these challenging cases. Until such developments emerge, clinical decision-making should prioritize established therapies with proven benefit, while carefully weighing the demonstrated risks of more aggressive interventional approaches.

## Data availability

The study protocol and additional supplementary information are available from the corresponding author upon reasonable request.

## Bibliography

1. Goyal M, Menon BK, van Zwam WH, et al. Endovascular thrombectomy after large-vessel ischaemic stroke: a meta-analysis of individual patient data from five randomised trials. Lancet Lond Engl. 2016;387(10029):1723–1731. doi:10.1016/S0140-6736(16)00163-X

2. Ospel JM, Goyal M. A review of endovascular treatment for medium vessel occlusion stroke. J Neurointerventional Surg. 2021;13(7):623–630. doi:10.1136/neurintsurg-2021-017321

3. Ospel JM, Menon BK, Demchuk AM, et al. Clinical Course of Acute Ischemic Stroke Due to Medium Vessel Occlusion With and Without Intravenous Alteplase Treatment. Stroke. 2020;51(11):3232–3240. doi:10.1161/STROKEAHA.120.030227

4. Menon BK, Hill MD, Davalos A, et al. Efficacy of endovascular thrombectomy in patients with M2 segment middle cerebral artery occlusions: meta-analysis of data from the HERMES Collaboration. J Neurointerventional Surg. 2019;11(11):1065–1069. doi:10.1136/neurintsurg-2018-014678

5. Thomalla G, Fiehler J, Subtil F, et al. Endovascular thrombectomy for acute ischaemic stroke with established large infarct (TENSION): 12-month outcomes of a multicentre, open-label, randomised trial. Lancet Neurol. 2024;23(9):883–892. doi:10.1016/S1474-4422(24)00278-3

6. Saber H, Desai SM, Haussen D, et al. Endovascular Therapy vs Medical Management for Patients With Acute Stroke With Medium Vessel Occlusion in the Anterior Circulation. JAMA Netw Open. 2022;5(10):e2238154. doi:10.1001/jamanetworkopen.2022.38154

7. Rizzo F, Romoli M, Simonetti L, et al. Reperfusion strategies in stroke with medium-to-distal vessel occlusion: a prospective observational study. Neurol Sci Off J Ital Neurol Soc Ital Soc Clin Neurophysiol. 2024;45(3):1129–1134. doi:10.1007/s10072-023-07089-w

8. Cascio Rizzo A, Schwarz G, Cervo A, et al. Safety and efficacy of endovascular thrombectomy for primary and secondary MeVO. J Stroke Cerebrovasc Dis Off J Natl Stroke Assoc. 2024;33(1):107492. doi:10.1016/j.jstrokecerebrovasdis.2023.107492

9. Psychogios M, Brehm A, Ribo M, et al. Endovascular Treatment for Stroke Due to Occlusion of Medium or Distal Vessels. N Engl J Med. 2025;392(14):1374–1384. doi:10.1056/NEJMoa2408954

10. Goyal M, Ospel JM, Ganesh A, et al. Endovascular Treatment of Stroke Due to Medium-Vessel Occlusion. N Engl J Med. 2025;392(14):1385–1395. doi:10.1056/NEJMoa2411668

11. Toni D, Lorenzano S, Puca E, Prencipe M. The SITS-MOST registry. Neurol Sci Off J Ital Neurol Soc Ital Soc Clin Neurophysiol. 2006;27 Suppl 3:S260–262. doi:10.1007/s10072-006-0632-9

12. Sun D, Liu R, Huo X, et al. Endovascular treatment for acute ischaemic stroke due to medium vessel occlusion: data from ANGEL-ACT registry. Stroke Vasc Neurol. 2022;8(1):51–58. doi:10.1136/svn-2022-001561

13. Zhang L, Su F, Zhang J, et al. Mechanical Thrombectomy for Treatment of Acute Cerebral Infarction due to Distal Medium Vessel Occlusions: A Retrospective Cohort Study. Brain Behav. 2024;14(11):e70119. doi:10.1002/brb3.70119

14. Aldaher B, Behera A, Morsi RZ, et al. Endovascular thrombectomy for distal medium vessel occlusions: A literature review. J Stroke Cerebrovasc Dis Off J Natl Stroke Assoc. 2025;34(1):108134. doi:10.1016/j.jstrokecerebrovasdis.2024.108134

15. Rennert RC, Wali AR, Steinberg JA, et al. Epidemiology, Natural History, and Clinical Presentation of Large Vessel Ischemic Stroke. Neurosurgery. 2019;85(Suppl_1):S4. doi:10.1093/neuros/nyz042

16. Association between brain imaging signs, early and late outcomes, and response to intravenous alteplase after acute ischaemic stroke in the third International Stroke Trial (IST-3): secondary analysis of a randomised controlled trial. Lancet Neurol. 2015;14(5):485–496. doi:10.1016/S1474-4422(15)00012-5

